# Development of a core outcome set on traditional Chinese medicine (COS-TCM) for rheumatic heart disease (RHD): study protocol

**DOI:** 10.1101/2022.01.19.22269522

**Authors:** Xiaodi Sheng, Chao Chen, Zhaochen Ji, Haiyin Hu, Mingyan Zhang, Hui Wang, Bo Pang, Dong Zhang, Junhua Zhang, Liping Guo

## Abstract

**Introduction:** Rheumatic heart disease (RHD) is an important cause of acquired heart disease in children and adolescents globally. Clinical trials on RHD treatment with traditional Chinese medicine (TCM) are gradually increasing in China. However, because clinical trials are heterogeneous and report outcomes selectively, similar studies cannot be merged and compared, complicating assessing the effectiveness and safety of TCM, diminishes the value of clinical trials, and results in a waste of research resources. Therefore, there is an urgent need to develop a core outcome set of traditional Chinese medicine for rheumatic heart disease (COS-TCM-RHD). This study will report the protocol development process for COS-TCM-RHD.

**Methods and analysis:** A multidisciplinary Steering Committee will lead the development of this protocol through four stages (1). Establishing a comprehensive and systematic outcomes checklist through a systematic review of previously published research, retrieval of clinical trial registration centers, patient’s semi-structured interviews, and clinician’s questionnaire surveys; (2). Screen stakeholder groups from various fields to participate in the Delphi survey; (3). Two rounds of e-Delphi surveys will be conducted to determine the outcomes of various concerned stakeholder groups; (4). Hold a face-to-face consensus meeting to develop the COS-TCM-RHD.

**Ethics and dissemination:** Ethical approval has been granted by the Tianjin university of Traditional Chinese Medicine Ethics Committee. The findings will be published in peer-reviewed journals and the website of Chinese Clinical Trials for Core Outcome Set.

**Trial registration:** This study protocol has been prospectively registered with the Core Outcome Measures in Effectiveness Trials (COMET):

http://www.comet-initiative.org/Studies/Details/1743.

**Strengths and limitations of this study:**

▪ This protocol is the first core outcomes set registered on the Core Outcome Measures in Effectiveness Trials (COMET) website for the treatment of rheumatic heart disease by Traditional Chinese medicine.
▪ This study is guided by the Core Outcome Set-STAndards for Development and Core Outcome Set-Standardized Protocol Items, with recommendations of the COMET.
▪ A multidisciplinary Steering Committee will supervise this research, and stakeholders from different fields including clinicians, patients, methodologists, and COS developers will be engaged.
▪ Systematic reviews, qualitative research (patient’s semi-structured interviews and clinician’s questionnaire surveys), Delphi surveys, and consensus meetings will be used for core outcome set development.
▪ Traditional Chinese medicine is mainly used in China. Thus, the geographical distribution of stakeholders will be a limitation.

## Background

Rheumatic heart disease (RHD) is a cardiac sequela of one or more episodes of rheumatic fever (RF), an autoimmune disease caused by group A streptococcus infection.^1^ The incidence of RHD is directly related to sanitary conditions.^2, 3^ In low and middle-income countries, it is the main cause of cardiovascular death in children and adolescents.^4^ In 2015,^5^ there were approximately 33.4 million patients with RHD worldwide, and 320,000 people died as a result. In 2019,^6^ the number of RHD cases globally was about 40 million, and approximately 340,000 people died. The number of patients with RHD is likely to continue to rise in the coming years.^7^ Therefore, RHD is still one of the major diseases affecting human health. ^8^

Western medicine (WM) treatment of RHD includes primary prevention (early detection and treatment of rheumatic fever), secondary prevention (application of antibiotics such as penicillin), and tertiary prevention (medical and surgical treatment of rheumatic heart disease and complications).^9, 10^ Although these measures can improve clinical symptoms to a certain extent, the treatment cost is expensive.^11^ Furthermore, long-term antibiotic therapy may result in major adverse effects, lowering the quality of life of RHD patients.

Traditional Chinese medicine (TCM) has rich experience and a unique theoretical system.^12^ In recent years, an increasing number of clinical trials of RHD on TCM have been published, confirming the important role of TCM treatment.^13-15^ We sorted out the outcomes of the randomized controlled trials (RCT) on RHD treatment with TCM. We found the following problems^[16]^: unclear definition of primary and secondary outcomes, a limited selection of surrogate outcomes, contempt of endpoint and safety outcomes, inconsistent measurement time points, etc. The issues mentioned above make it difficult to synthesize results in systematic reviews to do a meta-analysis.^17, 18^ The research findings cannot match the needs of relevant groups, and the efficacy cannot be accurately reflected, lowering the value of clinical trials and wasting research resources.^19, 20^

These issues can be addressed by developing a core outcome set (COS).^21^ Core Outcome Measures proposed the COS in Effectiveness Trials (COMET) initiative in 2010.^22^ COS is the minimum set of unified and standardized outcomes must be measured and reported in clinical trials of specific diseases.^23^ The development of COS aims to facilitate the comparison of results of similar clinical studies, but to reduce the risk of selective reporting,^24^ thus improving the quality and relevance of clinical studies and saving the study design cost.^25, 26^

The number of COS studies registered on COMET has increased significantly in recent years.^27^ But there is still no COS related to Chinese medicine intervention in RHD. As a result, a core outcome set of traditional Chinese medicines for rheumatic heart disease (COS-TCM-RHD) is required to address clinical research demands.

## Objective

The purpose of this study is to suggest a development strategy for the COS-TCM-RHD, this COS may be used for interventional research evaluating the therapeutic efficacy of Chinese medicine in RHD.

## Methods and analysis

### Steering Committee

A steering committee will be formed to examine and confirm research plans, provide advice, settle discrepancies throughout the research, and participate in consensus sessions. The steering committee will consist of eight experts, two cardiologists, two TCM physicians, two methodologists, one clinical researcher, and one COS developer. They will select one of them to serve as the committee leader.

### Working group

The main tasks of the working group consist of distributing questionnaires, statistical results, and holding meetings. The working group will be composed of 8 members, including one TCM clinician and one WM clinician, one methodologist, two professors, and three postgraduates from Chinese Clinical Trials COS Research Centre (ChiCOS), Tianjin University of Traditional Chinese Medicine (TJUTCM), China.

### Design

This development method of COS-TCM-RHD will reference the guidelines from the Core Outcome Set-STAndards for Development (COS-STAD),^28^ the Core Outcome Set-Standardized Protocol Items (COS-STAP),^29^ and the Core Outcome

Set-STAndards for Reporting (COS-STAR).^30^ This research will be conducted in four stages (**Figure 1**).

**Figure 1.**
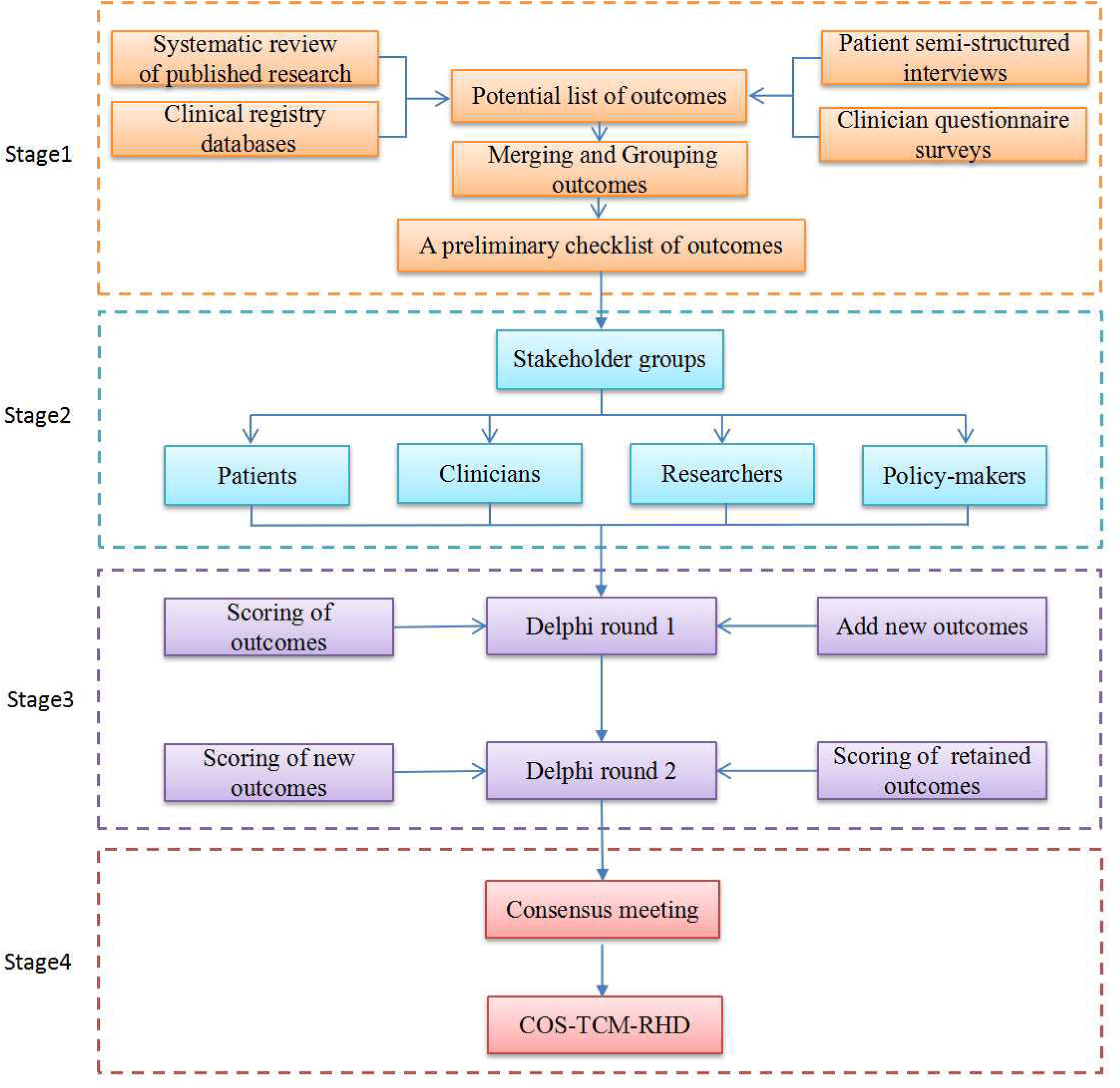
The development process for COS-TCM-RHD

Stage 1. Identifying potential outcomes, merging and grouping the outcomes to generate a checklist of preliminary outcomes entries.

Stage 2. Screening Stakeholder groups in various fields for the Delphi survey.

Stage 3. Conducting two rounds of e-Delphi surveys to determine the outcomes of the various stakeholder groups.

Stage 4. Holding a face-to-face consensus meeting to approve the final COS-TCM-RHD.

#### Stage 1: Identification of preliminary outcomes

First, a comprehensive outcomes pool will be established through searching databases and clinical trial registration centers in China and foreign countries, conducting patient’s semi-structured interviews and clinician’s questionnaire surveys; The working group will then standardize and categorize the outcomes and submit them for assessment to the steering committee to create a preliminary RHD outcome checklist.

##### Step 1. A systematic review of published research

###### Search Strategy

The following electronic databases will be searched: PubMed, Embase, Cochrane Library, the Web of Science (4 English databases), China National Knowledge Infrastructure, Wanfang Database, Chinese Biological Medicine Database, and the Chinese Scientific Journal Database (4 Chinese databases). The trials published from January 2011 to December 2021 will be included. The languages will be limited to English and Chinese. The search strategy of English databases is shown in Supplementary **File 1**.

###### Eligibility criteria

The inclusion and exclusion criteria for published articles will be as follows (see **Table 1**).

**Table 1.** Inclusion and exclusion criteria for published articles

| Items | Inclusion criteria | Exclusion criteria |
| --- | --- | --- |
| **Patients** | Patients diagnosed with rheumatic heart disease | Patients with other complications or acute exacerbations |
| **Intervention** | Measures related to TCM | None |
| **Outcome** | Outcomes included in all studies | None |
| **Study types** | Any type of studies, including RCT,case-control studies, cohort studies, etc | Studies with a primary aim of assessing the mechanisms or pharmacokinetics of interventions |
| **Language** | Chinese and English | Published in other languages |

###### Literature Selection

Two reviewers will examine the title and abstract independently and then the full text for another review. A third person will be consulted in the event of a disagreement. The PRISMA flowchart will be presented,^31^ including the number of selected and excluded studies and the reasons for exclusion.

###### Data extraction

The two researchers will independently extract and cross-check data. The extracted data including, (1). Basic information of the research, such as title, first author, s name, the time of publication, author’s area, etc. (2). Baseline characteristics of the research include age, course of the disease, sample size, TCM syndrome type (syndrome, symptoms, tongue, pulse), etc. (3). Intervention measures, including the name of intervention, course of treatment, frequency and dose of treatment, etc. (4). Outcome indicators, including outcome names or definitions, measurement methods, and time points of measurement.

##### Step 2. The search of clinical trial registries databases

###### Search Strategy

We’ll look at the clinical trials.gov and Chinese clinical trials registries. The search period will be from January 2011 to December 2021, with the keyword “rheumatic heart disease” as the search term.

###### Eligibility criteria

All clinical trial protocols for RHD with TCM will be included. The specific inclusion and exclusion criteria will be the same as the above-mentioned Systematic review.

###### Data Extraction

The extracted data will include the country of the registered organization/researcher, the status of the registered trial, Ethics approval, funding source, research stage, intervention measures, description of outcomes, outcome measurement methods, outcome measurement time points, etc.

##### Step 3. Patients semi-structured interviews

The patient’s opinion is important because the patient directly experiences the benefits and side effects of the treatment. In past COS investigations, semi-structured interviews were commonly employed to obtain patient perspectives.^32, 33^ Therefore, we will conduct semi-structured patient interviews to supplement patient concerns but not reported outcomes.

###### Eligibility criteria

RHD patients who are ≥18 years old and have experience in TCM treatment will be recruited from the First Affiliated Hospital of TJUTCM, the Second Affiliated Hospital of TJUTCM, Tianjin Academy of TCM Affiliated Hospital, and the Fourth Affiliated Hospital of TJUTCM. Patients with serious mental problems or communication difficulties and other factors that are not conducive to the development of the trial will be excluded.

###### Sampling

Referring to the experience of semi-structured interviews in previous COS research,^34, 35^ The sample size of patients in this study will be at least 40 cases. We will fully consider the patient’s gender, age, the stage of RHD, and the diversity of treatment history.

###### Protection of participants

The trained members of the workgroup will explain the purpose of the interview to patients and ask them whether they want to participate. Patients who agree to participate will sign the basic information form and informed consent form, and the patients will be informed that they can withdraw at any time.

###### Topic guide

The form of communication will be face-to-face, and interview records will be completely anonymized. First, the working group members will introduce the content of this study to the participants and then ask the patients to point out the outcomes that they thought were important through asking and guiding. The outline content is shown in **Table 2**. The outline content will be piloted and updated as needed.

**Table 2.** Questions about semi-structured interviews

| **Number** | **Questions** |
| --- | --- |
| 1 | When was the RHD diagnosed? |
| 2 | What troubles you most after RHD? |
| 3 | What degree of recovery do you want to achieve through treatment? |
| 4 | What are the outcomes you care about? Which is the most important? |

##### Step 4: Clinicians’ questionnaire survey

We will also use computerized questionnaires to acquire relevant outcomes by clinicians but not documented in the literature to diversify the sources of outcomes.

###### Eligibility criteria

The clinician should be specialized in TCM, WM, and integrated TCM and WM. Their main research direction will be cardiovascular disease, and they must have more than three years of medical experience.

###### Sampling size

Referring to previous similar COS research,^36, 37^ we expect to recruit 40 participants. To ensure that the survey is representative, we will recruit participants from five distinct areas and hospital levels (including outpatient and inpatient departments). The clinicians will be obtained from the membership lists of the China Integrated TCM and WM Professional Committee and the Chinese Society of Chinese Medicine Cardiovascular Disease Professional Committee.

###### Protection of participants

This is a volunteer study, physicians who participate must complete an informed consent form, a general information form (education, major, age, gender, and work unit), and an outcome questionnaire.

###### Topic guide

Since the doctor group is very familiar with clinical issues, to avoid the restriction of outcome selection, the questionnaire will adopt an open design without providing outcome options, allowing doctors to list important outcomes freely. Finally, the working group will summarize the survey information.

##### Step 5: Form a checklist of preliminary outcomes

The working group will summarize the data collected by the above four channels, conduct data analysis, and review it by the steering committee. If there is a difference, it will be resolved through discussion or consultation with a third party. The process will further comprise of four steps:

1. All outcomes will be numbered and imported into the Excel table, which is convenient to find the source.
2. Standardize the original retrieved outcomes by standardizing result names, visualizing and integrating outcomes, eliminating duplicate outcomes, and so on, before sorting and recording all outcome names and frequencies.
3. According to the functional attributes of the outcomes, the initial outcome items will be categorized into seven outcome domains^[38]^: symptoms and signs, TCM disease syndrome, quality of life, physical and chemical examination, economic evaluation, long-term prognosis, and adverse events.
4. The outcomes will be reviewed by the steering committee. If all outcomes exceed 100, the steering committee will vote internally to delete the outcomes. The outcomes approved by the steering committee will serve as the initial checklist of outcomes and will be included in the next Delphi survey.

#### Stage 2: Selection of Delphi stakeholder groups

The selection of stakeholders is crucial in the production of COS.^39, 40^ Referring to COS-STAD, we will recruit stakeholders from different fields and divide them into 4 groups: patients, clinicians, researchers, and policy-makers, to ensure that the views of each stakeholder are treated equally in the Delphi survey. The greater the number of stakeholder groups, the more reliable it may be,^41^ but too many people may also increase the difficulty of investigation and consensus. Considering that there is no consensus on the sample size of the Delphi survey,^42^ our Delphi will recruit 60 participants, 15 for each stakeholder group. We will send e-mails to more relevant stakeholders considering the response rate.

An e-invitation letter will be sent to the mailbox of the potential team member, explaining the content of this research and the importance of Delphi and stating that participants have the right to voluntarily, anonymously, and withdraw from the research at any time. Clicking on the link in the invitation letter means agreeing to participate in this research. Each member will be assigned a unique identification number to enable subsequent statistics and data storage.

**Group 1;** Patients ≥18 years of age diagnosed with RHD will be eligible for selection. We will post posters in the public areas of the hospital and will contact the doctors in the cardiology department via e-mail to invite them to participate in RHD patients who are undergoing or have already been diagnosed and treated.

**Group 2;** Clinicians, we will recruit clinicians through the hospital’s official website and WeChat public account, including cardiologists, TCM physicians, nurses, clinical pharmacists, etc. In addition, clinicians recommended by the steering committee will be accepted.

**Group 3;** Researchers, including methodological experts, clinical researchers, statistical experts, journal editors, COS developers, and other related experts. We will use social media for advertising job openings, and we will also screen high-impact RHD studies, contact the authors, and ask if they are interested in participating.

**Group 4;** Policy-makers, including health management personnel at various levels such as the country, province, city, county, etc. We will contact them by phone or e-mail and ask if they will participate in this survey.

#### Stage 3: Online Delphi surveys

Delphi survey is a group promotion technology that transforms individual opinions into group consensus through an iterative multi-stage process.^43^ We will conduct two rounds of Delphi surveys in electronic questionnaires, ask stakeholders to score outcomes, and reach a preliminary consensus on the outcomes for RHD treatment with TCM.

##### Scoring method

We will use the 9-point Likert-scaleto score candidate outcomes,^44, 45^ which was developed by the Grading of Recommendations Assessment, Development and Evaluation (GRADE) Working Group, and has been recommended by the COMET group^46^ to be widely used in the development of COS^47^ (see **Table 3**).

**Table 3.** Showing the 9-point Likert-scale

| **Score** | **Degree of importance** |
| --- | --- |
| 1-3 | not important for inclusion |
| 4-6 | important but not critical |
| 7-9 | critical for inclusion |

##### Round 1

###### Implementation Process

All attendees will be asked to fill out a basic information registration form. The basic information form will be developed following the features of various groupings. For example, the patient group will be asked to fill in disease information, current treatment plan, etc. and the doctor group will be asked to fill in the job title, position, department, etc. All candidate outcomes will be compiled and sent to all stakeholders via e-mail. Participants will be required to score every outcome on the Likert scale described above. To maintain the integrity of the outcomes, we will also invite participants to add other outcomes that they believe are important but are not on the list without scoring them. Participants will have three weeks to complete the questionnaire survey, and an e-mail reminder will be sent out at the ending of the second week.

###### Data Statistics and Analysis

Following the completion of the first round of the survey, members of the working group will collect all completed questionnaires, record the number of participants and responses, and compute the average score of each outcome and the score distribution of each stakeholder group. The steering committee will review any new recommended results. In the next eDelphi round, all of the saved results will be presented.

##### Round 2

###### Implementation Process

All participants completing round 1 will be invited to join round 2 of Delphi. The total number of participants, the proportion of each group and the entire group, the comparison of their scores with other group members, and new additional round 1 outcome will be summarized and presented to participants. Participants will be required to use the same scoring criteria as round 1 and re-scoring the retention outcomes that meet the requirements for entering round 2 (including the newly added outcomes in round 1). If any participant’s score changes, they will be asked to provide reasons. They will have three weeks to complete the online survey, and the working group will send an e-mail to participants at the ending of the second week reminding them to respond on time.

###### Data Statistics and Analysis

The working group will also count the number of participants and respondents, the average score of every outcome, and the distribution of scores for each stakeholder group. In addition, participants’ ratings from rounds 1 and 2 will be compared to assess potential risk deviations, and achieve”consensus in”, “consensus out” and “no consensus”. Statistically significant results will be discussed at the consensus meeting. The definition of consensus will be specified in advance, as shown in **Table 4**. Finally, the working group will compile all findings into a table and submit it for evaluation to the steering committee.

**Table 4.** Definition of consensus

| **Classification** | **Definition** |
| --- | --- |
| Consensus in | $\geq 70\%$ participants scoring 7 to 9 and $< 15\%$ participants scoring 1 to 3 |
| Consensus out | $\geq 70\%$ participants scoring 1 to 3 and $< 15\%$ participants scoring 7 to 9 |
| No consensus | Anything else |

#### Stage 4: Consensus meeting

In the last step of this study, a face-to-face consensus meeting will be held to discuss and review the final COS. The conference will be held in Tianjin, China, for one day. This meeting will be open to all members of the steering committee. Participants who have completed both rounds of the Delphi survey are also eligible to represent. To reduce the imbalance in stakeholder representation, we will select participant representatives from each stakeholder group at random. The overall number of attendees for this meeting is expected to be 35. The meeting will be hosted by an experienced moderator who will have no voting privileges.

##### Consensus meeting Content

The working group will present the results of the second round of e-Delphi surveys to the participants. According to **Table 4**, the “Consensus in” outcomes are immediately included in the final COS, whereas the “Consensus out” outcomes are directly eliminated. Participants will vote anonymously on the “No consensus” outcomes to determine whether it is “Consensus in” or “Consensus out.” If there is any conflict of view, it will be resolved by the steering committee through the Modified Nominal Group Technique (mNGT).^48^ After the consensus meeting, the working group will sort out the meeting minutes and form the final COS-TCM-RHD.

## Discussion

There are some limitations in the outcomes presented in TCM for RHD clinical trials, making it very difficult to combine and evaluate similar research and complicating therapeutic decision-making. This research program is the first COS-TCM-RHD registered at COMET. Our study plan fully adopted the views of multiple stakeholder groups through Systematic review, semi-structured interviews, Clinicians’ questionnaires, e-Delphi surveys, and consensus meetings, which is planned to develop a COS-TCM-RHD with high feasibility and promotion.

After the final COS is constituted, we will agree on how and when to measure outcomes. Therefore, future work will conclude the evaluation of core outcomes measurement instruments following consensus-based standards for selecting health measurement instruments for the core outcomes.^49^ This will include systems review, quality assessment of measurement tools, and consensus among stakeholder groups.^50, 51^

The COS is widely used in clinical and scientific research, thus it will help increase the consistency of research, reduce the heterogeneity of reports, and improve the value of clinical trials of TCM. COS can improve the clinical care and patients’ experience when it is relevant to both clinicians and patients. Furthermore, COS could help healthcare providers to better understand the needs of patients and adjust funding priorities for interventions. In conclusion, we expect that the COS-TCM-RHD will play a pivotal role in improving the quality of clinical trials, patient care, clinical decision-making, and policymaking.

### Ethics and dissemination

The Ethics Committee has approved the entire project of the Tianjin University of Traditional Chinese Medicine Research Ethics Committee (TJUTCM-EC20210008). We will obtain informed consent from all participants who participate in semi-structured interviews, Delphi surveys, and consensus meetings.

After completing the final COS, we will publish the findings in peer-reviewed and open access journals, report them at national and international meetings, and disseminate them on the ChiCOS (www.ChiCOS.org.cn). We also intend to send the publication to all participants of this study. It is hoped that the scope of COS publicity can be expanded and recognized by relevant industry associations.

## Data Availability

All data produced in the present study are available upon reasonable request to the author

## Abbreviations

RHD: Rheumatic heart disease
RF: Rheumatic fever
WM: Western medicine
TCM: Traditional Chinese Medicine
RCT: Randomized controlled trials
COS: Core Outcome Set
COMET: Core Outcome Measures in Effectiveness Trials
COS-TCM-RHD: Core outcome set of traditional Chinese medicine for rheumatic heart disease
CHICOS: Chinese Clinical Trials COS Research Centre
TJUTCM: Tianjin University of Traditional Chinese Medicine
COS-STAR: Core Outcome Set- STAndards for Reporting
COS-STAD: Core Outcome Set- STAndards for Development
COS-STAP: Core Outcome Set- Standardized Protocol Items
GRADE: Grading of Recommendations Assessment, Development, and Evaluation
mNGT: Modified Nominal Group Technique

## Contributions

XS and CC wrote the paper and conceived the project. XS and CC contributed equally to the manuscript. CC contributed to the English writing and translation of this protocol. ZJ and HH are responsible for conducting the systematic review. HW, BP, MZ, and DZ participated in the design of the study. LG and JZ provide supervision for the project. All authors edited and critically revised the study protocol. All authors have read, contributed, and approved the manuscript.

## Patient consent for publication

Not required.

## Provenance and peer review

Not commissioned; externally peer reviewed.

