## Supplementary File 1 for "Development of a core outcome set on traditional Chinese medicine (COS-TCM) for rheumatic heart disease (RHD): study protocol"

### The Detailed Search Strategy

**The Pubmed database Search Strategy**

**#1** ("Rheumatic heart disease"[MeSH Terms]) OR ("Rheumatic heart disease"[Title/Abstract] OR "Rheumatic Heart Diseases"[Title/Abstract] OR "Bouillaud Disease"[Title/Abstract] OR "Bouillaud's Disease"[Title/Abstract] OR "Rheumatic valve disease"[Title/Abstract] OR "Rheumatic heart-failure"[Title/Abstract] OR "Rheumatic cardiac disease"[Title/Abstract] OR "Chronic rheumatic heart disease"[Title/Abstract] OR "Rheumatic mitral valvular disease"[Title/Abstract] OR "Rheumatic cardiopathy"[Title/Abstract] OR "Rheumatoid heart disease"[Title/Abstract] OR "Rheumatic heart valve disease"[Title/Abstract] OR "Rheumatic valvular heart disease"[Title/Abstract] OR "Rheumatic valvular disease"[Title/Abstract])

**#2** (("medicine, chinese traditional"[MeSH Terms]) OR (Chinese herbal drugs[MeSH Terms])) OR ("Traditional Chinese Medicine"[Title/Abstract] OR TCM[Title/Abstract] OR "Chinese herbal drugs"[Title/Abstract] OR "Chinese patent drugs"[Title/Abstract] OR "integrated TCM WM"[Title/Abstract])

**#3** #1 AND #2

**The Cochrane library database Search Strategy**

**#1** ("Rheumatic heart disease" OR "Rheumatic Heart Diseases" OR "Bouillaud Disease" OR "Bouillaud's Disease" OR "Rheumatic valve disease" OR "Rheumatic heart-failure" OR "Rheumatic cardiac disease" OR "Chronic rheumatic heart disease" OR "Rheumatic mitral valvular disease" OR "Rheumatic cardiopathy" OR "Rheumatoid heart disease" OR "Rheumatic heart valve disease" OR "Rheumatic valvular heart disease" OR "Rheumatic valvular disease"):ti,ab,kw

**#2** ("Traditional Chinese Medicine" OR TCM OR "Chinese herbal drugs" OR "Chinese patent drugs" OR "integrated traditional Chinese medicine and western medicine" OR "integrated TCM WM"):ti,ab,kw

**#3 #1** AND **#2**

**The Embase database Search Strategy**

**#1** 'rheumatic heart disease':ab,ti OR 'rheumatic heart diseases':ab,ti OR 'bouillaud disease':ab,ti OR 'bouillauds disease':ab,ti OR 'rheumatic valve disease':ab,ti OR 'rheumatic heart-failure':ab,ti OR 'rheumatic cardiac disease':ab,ti OR 'chronic rheumatic heart disease':ab,ti OR 'rheumatic cardiol disease':ab,ti OR 'rheumatic mitral valvular disease':ab,ti OR 'rheumatic cardiopathy':ab,ti OR 'rheumatoid heart disease':ab,ti OR 'rheumatic heart valve disease':ab,ti OR 'rheumatic valvular heart disease':ab,ti OR 'rheumatic valvular disease':ab,ti

**#2** 'traditional chinese medicine':ab,ti OR TCM:ab,ti OR 'chinese herbal drugs':ab,ti OR 'chinese patent drugs':ab,ti OR 'integrated traditional chinese medicine AND western medicine':ab,ti OR 'integrated tcm wm':ab,ti

**#3** #1 AND #2

**Web of Science**

**#1** 'rheumatic heart disease':ab,ti OR 'rheumatic heart diseases':ab,ti OR 'bouillaud disease':ab,ti OR 'bouillauds disease':ab,ti OR 'rheumatic valve disease':ab,ti OR 'rheumatic heart-failure':ab,ti OR 'rheumatic cardiac disease':ab,ti OR 'chronic rheumatic heart disease':ab,ti OR 'rheumatic cardiol disease':ab,ti OR 'rheumatic mitral valvular disease':ab,ti OR 'rheumatic cardiopathy':ab,ti OR 'rheumatoid heart disease':ab,ti OR 'rheumatic heart valve disease':ab,ti OR 'rheumatic valvular heart disease':ab,ti OR 'rheumatic valvular disease':ab,ti

**#2** 'traditional chinese medicine':ab,ti OR TCM:ab,ti OR 'chinese herbal drugs':ab,ti OR 'chinese patent drugs':ab,ti OR 'integrated traditional chinese medicine AND western medicine':ab,ti OR 'integrated tcm wm':ab,ti

**#3** #1 AND #2
